# Trans-ethnic analysis reveals genetic and non-genetic associations with COVID-19 susceptibility and severity

**DOI:** 10.1101/2020.09.04.20188318

**Authors:** Janie F. Shelton, Anjali J. Shastri, Chelsea Ye, Catherine H. Weldon, Teresa Filshtein-Somnez, Daniella Coker, Antony Symons, Jorge Esparza-Gordillo, Team The 23andMe COVID-19 Team, Stella Aslibekyan, Adam Auton

## Abstract

COVID-19 presents with a wide range of severity, from asymptomatic in some individuals to fatal in others. Based on a study of over one million 23andMe research participants, we report genetic and non-genetic associations with testing positive for COVID-19, respiratory symptoms, and hospitalization. Risk factors for hospitalization include advancing age, male sex, elevated body mass index, lower socio-economic status, non-European ancestry, and pre-existing cardio-metabolic and respiratory conditions. Using trans-ethnic genome-wide association studies, we identify a strong association between blood type and COVID-19 diagnosis, as well as a gene-rich locus on chr3p21.31 that is more strongly associated with outcome severity. While non-European ancestry was found to be a significant risk factor for hospitalization after adjusting for socio-demographics and pre-existing health conditions, we did not find evidence that these two primary genetic associations explain differences between populations in terms of risk for severe COVID-19 outcomes.

## Introduction

The COVID-19 pandemic has caused unprecedented disruption to modern societies throughout the world. Since the emergence of the disease, it has become clear that the course of disease can vary considerably between individuals^1^, with some experiencing mild or non-existent symptoms and others experiencing severe outcomes, including hospitalization or even death. It has been well documented that a number of host factors are correlated with disease progression, with primary risk factors including sex, age, ethnicity, and the presence of underlying medical conditions^2^.

Much less is known about the genetic basis of COVID-19 disease risk, both in terms of susceptibility to infection and severity of outcomes following infection. It has been well-established that genetics plays a role in host susceptibility to infection and disease pathogenesis in humans^3^. Notable examples include the protective effects of the *CCR5Δ32* mutation on infection with the *HIV-1* virus^4^ and the sickle-cell mutation in the *HBB* gene offering protection against the malaria-causing *Plasmodium falciparum*^5^. Over the past decade, genome-wide association studies (GWAS) have proved to be a useful tool for uncovering novel infectious disease susceptibility loci, identifying loci associated with pathogen clearance or persistence, and providing supporting evidence for the role of certain host factors implicated in disease progression and severity^6,7^.

Given the rapid emergence of COVID-19, pre-existing genetic cohorts offer a path to rapid data collection that can address questions surrounding the relationship between host genetics and COVID-19 in a timely fashion. Amongst the largest pre-existing genetic cohorts are those that have been developed via direct-to-consumer (DTC) genetic testing. 23andMe is a DTC genetic testing company with over 10 million genotyped customers. As part of the 23andMe service, customers are genotyped on SNP microarrays and offered the opportunity to participate in scientific research, and approximately 80% of customers consent to do so. In general, research participation is conducted via online surveys, which research participants can complete at any time. Research participants are re-contactable and can be invited to participate in new surveys that are developed over time.

In this paper, we describe the engagement of the 23andMe research cohort to address questions surrounding COVID-19 risk factors and host genetics. Having collected data from over one million research participants, we identified 15,434 individuals who reported a positive COVID-19 test, of whom 1,131 reported hospitalization with COVID-19 symptoms. We first investigated non-genetic risk factors associated with COVID-19 severity and found that lower socio-economic status, African American ancestry, obesity, and pre-existing conditions associated with a higher risk of hospitalization. We subsequently conducted a GWAS of phenotypes related to both COVID-19 diagnosis and severity. We performed GWAS separately in samples of European, Latino, and African American ancestries and used the resulting data to perform a trans-ethnic meta-analysis. We identified a strong association with the *ABO* gene, which appears to be connected with COVID-19 diagnosis, and another strong association within a gene-rich locus at chr3p21.31, which appears to be connected with COVID-19 severity.

## Methods

### Overview of study recruitment and data collection

Participants in this study were recruited from the customer base of 23andMe, Inc., a personal genetics company. All individuals included in the analyses provided informed consent and answered surveys online according to our human subjects research protocol, which was reviewed and approved by Ethical and Independent Review Services, a private institutional review board (http://www.eandireview.com).

Primary recruitment was carried out by email to approximately 6.7 million 23andMe research participants over 18 years of age and living in the United States or the United Kingdom. Additionally, pre-existing customers were invited to participate in the study through promotional materials on the 23andMe website, the 23andMe mobile application, and via social media.

Study participation consisted solely of web-based surveys, including an initial baseline survey, and three follow-up surveys fielded 1-month following completion of the baseline survey. Because enrollment is ongoing, not all participants would have received, or completed all of the follow-up surveys. All (4) surveys included questions about symptoms of cold or flu-like illnesses from February of 2020 onward, COVID-19 diagnosis and testing, hospitalization, severity of illness, COVID-19 diagnosis of first and second degree family members, and potential sources of exposure to COVID-19. Other respondent characteristics, such as age, sex, pre-existing conditions, educational attainment, zip code, and smoking status had been collected via previously deployed surveys for the majority of participants, but were also queried in the COVID-19 baseline survey if the data were missing.

Due to the geographically localized nature of the COVID-19 outbreak during the study period, we geo-targeted the email recruitment campaign to follow the outbreak as it moved through the United States (Figure 1). Emails to each state/country were batched into tranches on the basis of the anticipated timing of the hospitalization demand peak within each region, as assessed from the Institute for Health Metrics and Evaluation (IHME) prediction model^8^. Each tranche was recruited via email a minimum of two weeks after estimated peak hospitalization demand, as determined by the IHME predictive models. IHME predictions varied over the course of the study, and the order in which regions were targeted was adjusted accordingly. The email send dates are detailed in Supplementary Table 1.

**Figure 1:**
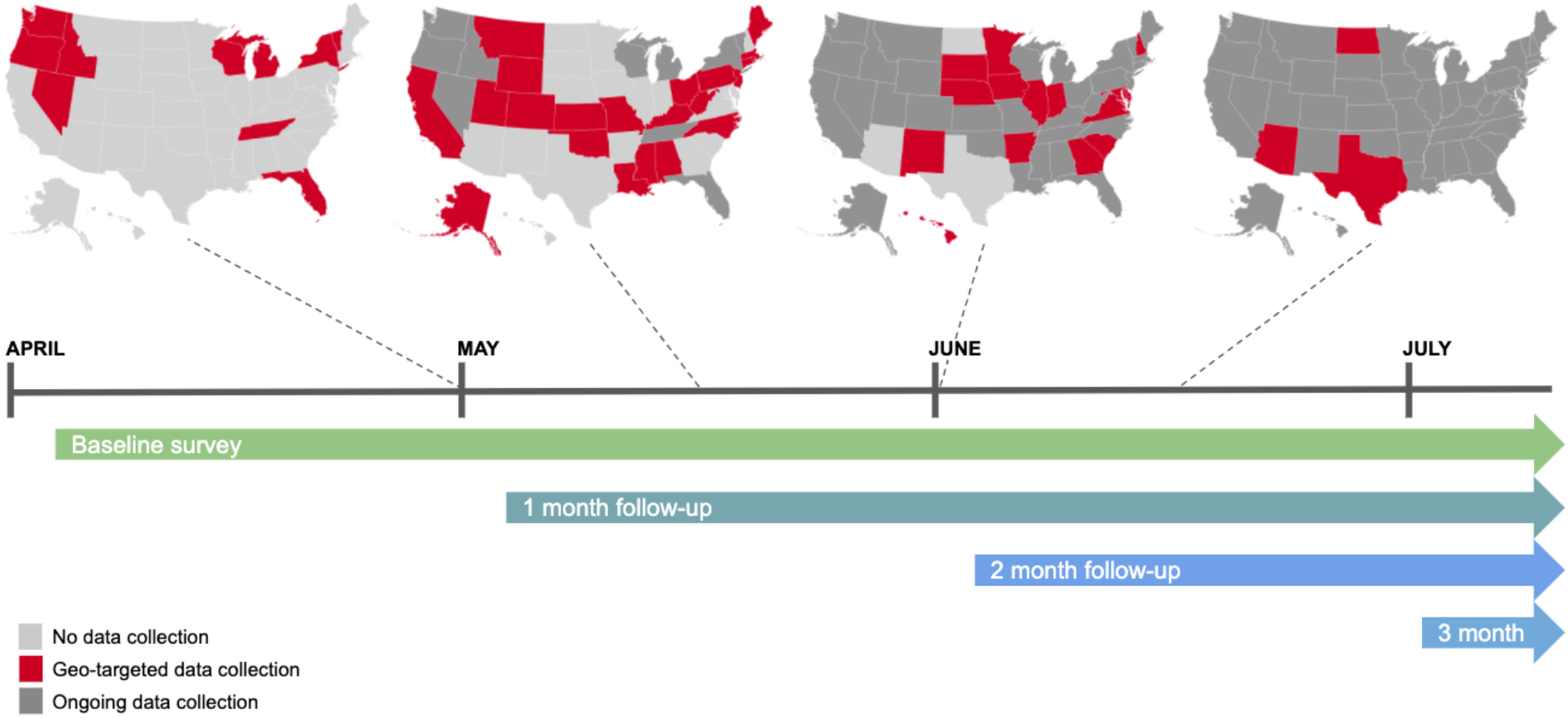
COVID-19 email-based study recruitment in the United States between 6 April and 25 July 2020.

To visualize the estimated prevalence of test positive COVID-19 study participants across the United States, we calculated the proportion of test positive cases among participants for each county with greater than 10 responses. In order to generate a smoothed representation of the data, we identified the 15 nearest counties for each county, and calculated a prevalence estimate weighted by sample size across those counties. To highlight how the smoothed version represents the raw data, we also generated a version without smoothing, with a random 10% of study participants removed in order to eliminate the possibility of reidentification in the dataset.

### Social, demographic, and pre-existing conditions evaluated as risk factors for COVID-19 hospitalization

To explore non-genetic factors associated with hospitalization for COVID-19, age, sex, ancestry, median household income of the residential zip code, educational attainment, body mass index, and pre-existing conditions were evaluated as risk factors in bivariate and multivariate logistic regression models. Ancestry was inferred via a previously described genetic ancestry classification algorithm^9,10^ to ensure compatibility with the GWAS methodology described below. As social and demographic factors are strongly associated with COVID-19 hospitalization and many of the pre-existing conditions, final models estimating the relationship between the preexisting condition and COVID-19 hospitalization were adjusted for age (10-year increments), sex, ancestry, high school or below education, and median household income of the residential zip code (in $10,000 increments). Finally, a multivariate logistic regression model was specified to quantify the risk of the top conditions after adjusting for each other, and for sociodemographic characteristics related to risk of hospitalization for COVID-19.

The relationship between age and COVID-19 and hospitalization was determined by categorizing cases by 10-year age increments between 30 and 80, and then calculating the percentage of cases in each age group and the percentage of cases which reported hospitalization. To describe differences in hospitalization by ancestry, age-standardized estimates were calculated by applying the percent of cases hospitalized within age strata for European, African American, and Latino respondents and applying that percentage to the age structure of all cases in the study population. While our data also included respondents of other ancestries, such as East Asian and South Asian, the sample sizes for these populations were too small for robust inferences to be performed. All analyses were conducted in R statistical software, version 3.3.

### Phenotype definitions for GWAS

Using the information derived from the surveys, we defined a set of phenotypes that aimed to capture aspects of COVID-19 diagnosis and severity. Following preliminary analysis, we selected one ‘diagnosis’ phenotype that contrasts positive and negative outcomes from a COVID-19 test, and four phenotypes that capture aspects of COVID-19 disease ‘severity’, including pneumonia, hospitalization, or the need for respiratory support (Supplementary Table 2). For the severity phenotypes, we explored different choices of how to define controls, and found that using controls who had not reported a COVID-19 diagnosis (were neither diagnosed with nor tested positive for COVID-19) provided a large-scale control set that appeared to maximize power in the GWAS.

### Genotyping and SNP imputation

DNA extraction and genotyping were performed on saliva samples by CLIA-certified and CAP-accredited clinical laboratories of Laboratory Corporation of America. Samples were genotyped on one of five genotyping platforms. The V1 and V2 platforms were variants of the Illumina HumanHap550 + BeadChip and contained a total of about 560,000 SNPs, including about 25,000 custom SNPs selected by 23andMe. The V3 platform was based on the Illumina OmniExpress + BeadChip and contained a total of about 950,000 SNPs and custom content to improve the overlap with our V2 array. The V4 platform was a fully custom array of about 950,000 SNPs and included a lower redundancy subset of V2 and V3 SNPs with additional coverage of lower-frequency coding variation. The V5 platform was based on the Illumina GSA array, consisting of approximately 654,000 pre-selected SNPs and approximately 50,000 custom content variants. Samples that failed to reach 98.5% call rate were re-analyzed. Individuals whose analyses failed repeatedly were re-contacted by 23andMe customer service to provide additional samples, as is done for all 23andMe customers.

Participant genotype data were imputed using the Haplotype Reference Consortium (HRC) panel^11^, augmented by the Phase 3 1000 Genomes Project panel^12^ for variants not present in HRC. We phased and imputed data for each genotyping platform separately. For the non-pseudoautosomal region of the X chromosome, males and females were phased together in segments, treating the males as already phased; the pseudoautosomal regions were phased separately. We then imputed males and females together, treating males as homozygous pseudo-diploids for the non-pseudoautosomal region.

### Genome-wide association study (GWAS)

Genotyped participants were included in GWAS analyses on the basis of ancestry as determined by a genetic ancestry classification algorithm^10^. For each phenotype, we selected a set of unrelated individuals so that no two individuals shared more than 700cM of DNA identical by descent (IBD). For case-control phenotypes, if a case and a control were identified as having at least 700cM of DNA IBD, we preferentially discarded the control from the sample.

For case-control comparisons, we tested for association using logistic regression, assuming additive allelic effects. For tests using imputed data, we use the imputed dosages rather than best-guess genotypes. We included covariates for age, age squared, sex, a sex:age interaction, the top ten principal components to account for residual population structure, and dummy variables to account for genotyping platform. The association test p-value was computed using a likelihood ratio test, which in our experience is better behaved than a Wald test on the regression coefficient. Results for the X chromosome were computed similarly, with men coded as if they were homozygous diploid for the observed allele.

We ran GWAS for each phenotype separately, and combined both genotyped and imputed data. When choosing between imputed and genotyped GWAS results, we favored the imputed result, unless the imputed variant was unavailable or failed quality control (QC). For imputed variants, we removed variants with low imputation quality (r^2^ < 0.5 averaged across batches, or a minimum r^2^ < 0.3) or with evidence of batch effects (ANOVA F-test across batches, p-value < 10^−50^). For genotyped variants, we removed variants only present on our V1 or V2 arrays (due to small sample size) that failed a Mendelian transmission test in trios (p-value < 10^−20^), that failed a Hardy-Weinberg test in Europeans (p-value < 10^−20^), failed a batch effect test (ANOVA p-value < 10^−50^), or had a call rate < 90%.

We repeated the GWAS analysis separately in each population cohort for which we had sufficient data (European, Latino, African American), and then performed trans-ethnic meta-analysis using a fixed effects model (inverse variance method^13^), restricting to variants of at least 1% minor allele frequency.

Within each GWAS, we identified regions with genome-wide significant (GWS) associations. We define the region boundaries by identifying all SNPs with p-value < 10^−5^ within the vicinity of a GWS association, and then grouping these regions into intervals so that no two regions are separated by less than 250 kb. We consider the SNP with the smallest p-value within each interval to be the index SNP. We also annotated our findings based on linkage disequilibrium (LD) with results from published GWAS, coding variation, and expression quantitative trait loci (eQTL), specifically by finding annotations with r^2^ > 0.5 and within 500kb of the index SNP.

### Blood group analyses

We classified haplotypes into blood groups on the basis of genotypes at three SNPs: rs8176747, rs41302905, and rs8176719^14,15^. A deletion at rs8176719 confers a type O haplotype, as does a T allele at rs41302905. If neither rs8176719 nor rs41302905 confers type O, then rs8176747 distinguishes between types A and B. This assignment paradigm is described in Supplementary Table 3.

Given haplotype assignments, individuals were assigned a blood type on the basis of their diploid combination of haplotypes, with type O being recessive, so that individuals with two O haplotypes were assigned type O, individuals with one O and one A haplotype were assigned type A, and so on.

We note that the blood group assignment methodology described above is incomplete, and there are other rare variants that can influence blood group^14^. In order to understand the accuracy of the genetic blood group assignments, we compared to self-reported blood groups from over 1.47 million research participants. We found that the genetic assignments achieved 90-94% precision and 72-96% recall compared to the self-reported data, depending on blood group (Supplementary Table 4).

We tested for association between ABO blood group and COVID-19 phenotypes in each population using logistic regression, testing blood group pairs separately (e.g. individuals with blood group O vs individuals with blood group A) and only testing unrelated individuals. We included covariates for age, age squared, sex, a sex:age interaction, and the top ten principal components. We meta-analysed across populations using a fixed effects model. We repeated these analyses using self-reported blood group assignments in place of genetically determined assignments and found the results to be qualitatively similar (data not shown).

We performed a similar analysis between ABO blood groups and experience of influenza by considering research participants who answered the question “Have you had influenza (flu) in the past 12-months? *Common symptoms of flu are fever over 100° F (38° C), muscle aches, chills and sweats, headache, dry cough, fatigue, nasal congestion and sore throat. As compared to the common cold, symptom onset for influenza is faster, more severe, and can last 1-2 weeks*.” In order to avoid overlap with individuals reporting experiences with COVID-19, we tested for association between influenza and the ABO blood groups using a sample of individuals that answered the question during either the 2017-2018 flu season, defined as starting in October 2017 and ending in September 2018, or the 2018-2019 flu season, defined as starting in October 2018 and ending in September 2019.

To test for differences between rhesus positive and negative blood groups, we use the structural variant esv3585521 to obtain rhesus type. This variant, located within the *RHD* gene, has a 39.4% frequency in European populations and associates very strongly with self-reported rhesus type in 23andMe data (OR = 22.1, p-value = 1.8e-298). We take individuals imputed as homozygous for the deletion as being rhesus negative. Within each blood group, we tested for association between rhesus type and COVID-19 phenotypes in the European-ancestry population using logistic regression. We included covariates for age, age squared, sex, a sex:age interaction, and the top ten principal components.

## Results

### Respondent characteristics

As of July 25th 2020, just over 1.05 million research participants took the COVID-19 baseline survey. Respondents were included in this analysis if they had consented to research and had a non-missing response to the question, “*Have you been tested for COVID-19?*”. Of those, 15,434 self-reported a positive test result, and 1,131 reported hospitalization with a positive test (Table 1). The majority of respondents were currently based in the United States (93.2%), followed by the United Kingdom (2.4%), with the remainder responding from other countries around the world (4.4%). The majority of respondents were of European ancestry (80.3%), although the study also included substantial representation from Latino (11.3%, n = 118, 787), and African American or Black (2.7%, n = 28,592; hereafter referred to as African American) ancestries. Study participants were 63% female with a median age of 51 years.

Those reporting a positive test were more likely to be male (OR = 1.22, 95% CI 1.18-1.26, pvalue < 2.2e-16), younger on average (43.0 vs. 51.0, p-value < 0.001), and less likely to be of European ancestry (70.3% COVID-19 positive test were European vs. 80.3% of all study participants, p-value < 0.001), compared to other survey respondents. Comparing those who tested positive to all other study participants, living in an urban environment (90.5% of respondents vs. 95% of those with a COVID-19 positive test, p-value < 0.001) and employment as a healthcare professional (9.2% of respondents but 21.7% of COVID-19 positive tests, pvalue < 0.001) were associated with a higher likelihood of reporting a positive test. Pre-existing conditions were negatively associated with reporting a COVID-19 positive test, as was current smoking status (Table 1).

**Table 1:**
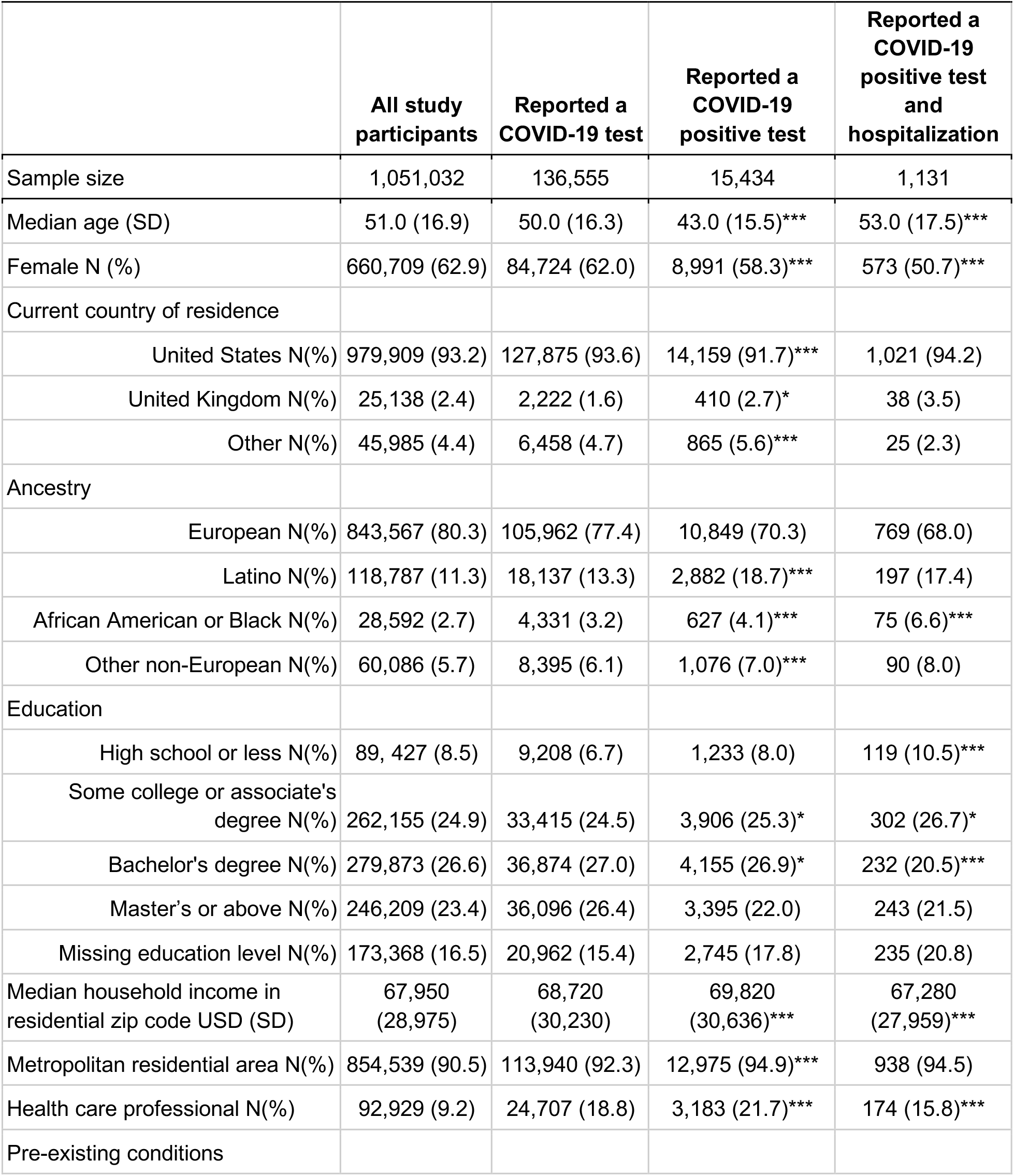
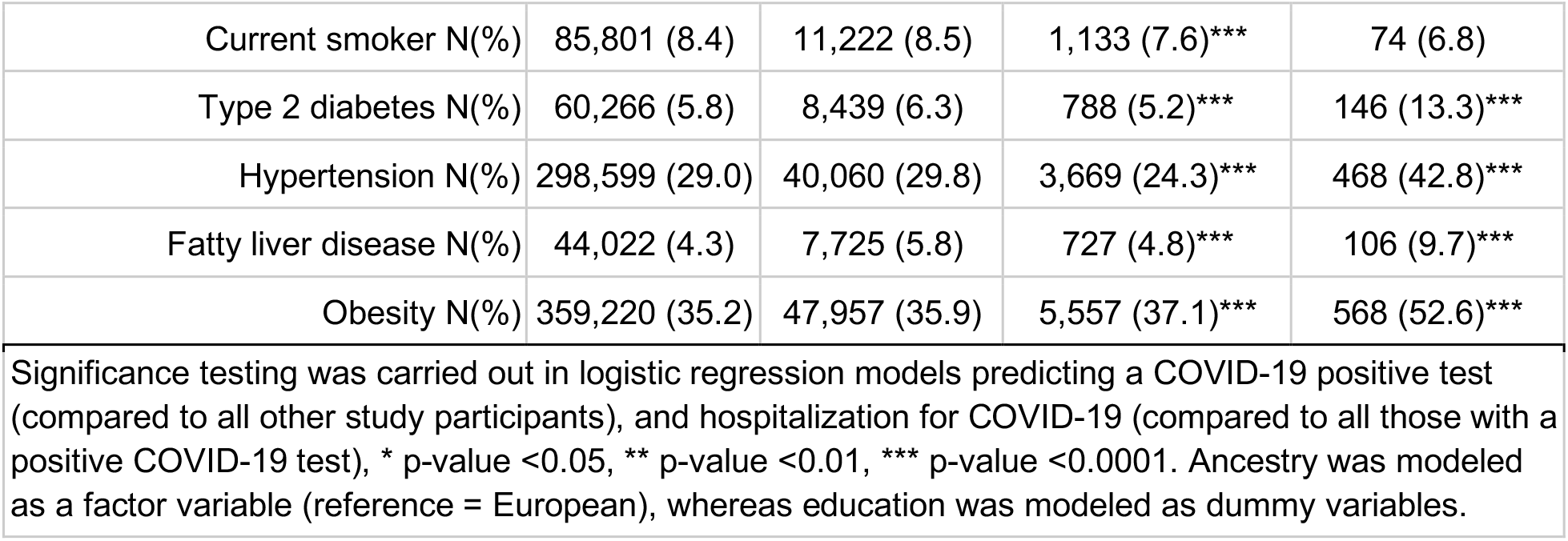
Demographic and health characteristics of COVID-19 survey respondents.

In addition to being more likely to report a positive test (1.7% vs. 1.4%, p-value < 2.2e-16; chi-squared test), male respondents were more likely to report hospitalization (10.1% vs. 7.4%) than female respondents (p-value = 4.3e-8; chi-squared test). While the proportion of individuals reporting a positive test declined as a function of age, hospitalization rates increase dramatically with age (Figure 2a). Generally, non-European ancestry was associated with higher rates of self-reported COVID-19 infection and higher proportions of hospitalization. For Latinos, the higher proportion of hospitalization was consistent with a higher proportion of individuals reporting a positive COVID-19 test as compared to other groups (O/E = 0.93, p-value = 0.27; chi-square test). However, for African Americans, the proportion reporting hospitalization was almost twice as high as expected from the proportion reporting a positive COVID-19 test (O/E = 1.96, p-value = 3e-11; chi-square test), implying either more severe outcomes for those that became infected or an under reporting of test positive status (Figure 2b).

**Figure 2:**
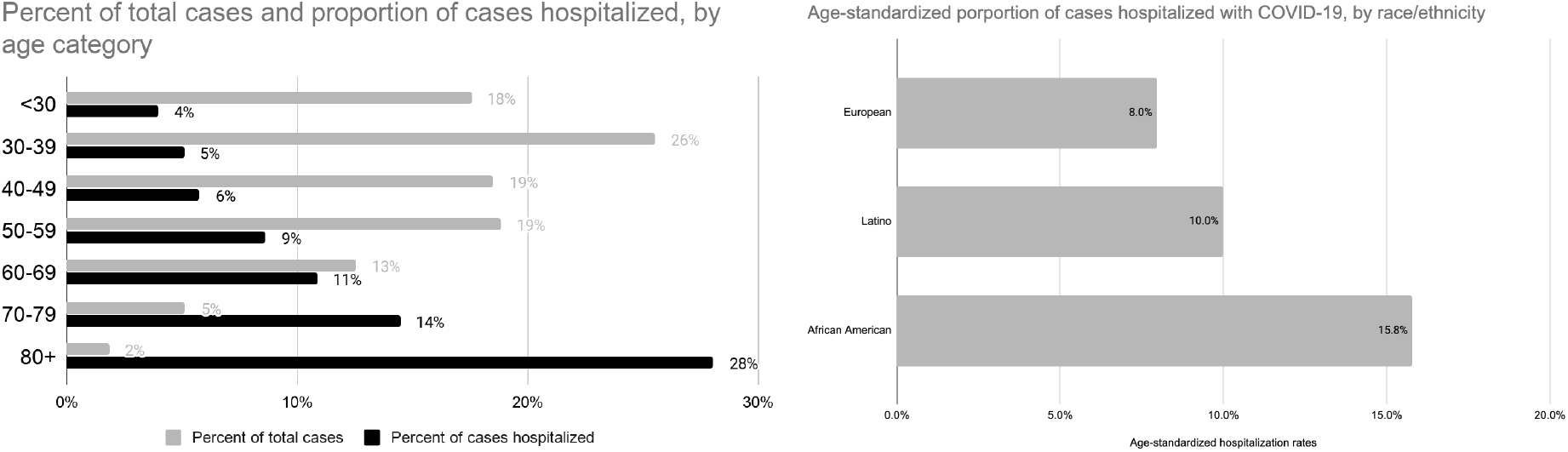
a) Distribution of COVID-19 cases and percent of total cases hospitalized by age. b) Age-standardized percent of cases reporting hospitalization by ancestry.

In models taking into account age, sex, education, ancestry, education level, and median household income in the residential zip code, among those with a COVID-19 positive test, several cardiometabolic and respiratory conditions were associated with an elevated risk of hospitalization. However, the strongest association was with obesity, which conferred a 2.5 fold increased risk of hospitalization (aOR = 2.45, 95% CI: 1.99-3.02). Type 2 diabetes, cardiovascular disease, hypertension, respiratory conditions including asthma, fatty liver disease, and gastro-esophageal reflux disease (GERD) were all positively associated with COVID-19 hospitalization, in models adjusting for BMI (Table 2).

**Table 2:** Adjusted odds ratios and 95% confidence intervals estimating the association between pre-existing conditions and hospitalization with COVID-19.

| Pre-existing condition | COVID-19 hospitalization |  |
| --- | --- | --- |
|  | Adjusted OR | 95% CI |
| Body Mass Index |  |  |
| Underweight | 1.30 | 0.81 - 2.09 |
| Normal weight | 1.00 | REF |
| Overweight | 1.34 | 1.07 - 1.68*** |
| Obese | 2.45 | 1.99 - 3.02*** |
| Other cardio-metabolic |  |  |
| Type 2 diabetes | 1.66 | 1.30 - 2.11*** |
| Any cardiovascular disease | 1.35 | 1.15 - 1.60*** |
| Coronary artery disease | 1.48 | 1.02 - 2.14* |
| Arrhythmia | 1.49 | 1.22 - 1.83*** |
| Hypertension | 1.40 | 1.18 - 1.66*** |
| Renal/hepatic |  |  |
| Chronic kidney disease | 1.35 | 0.94 - 1.94 |
| Fatty liver disease, including NASH | 1.76 | 1.36 - 2.28*** |
| Hepatitis | 0.89 | 0.60 - 1.33 |
| Respiratory |  |  |
| Any underlying lung or respiratory condition | 1.44 | 1.21 - 1.72*** |
| COPD | 1.35 | 0.88 - 2.07 |
| Asthma | 1.27 | 1.07 - 1.51** |
| Other |  |  |
| GERD | 1.24 | 1.06 - 1.45** |
| All models were adjusted for sex, age (10 yr increments), education level, BMI (categorical), genetic ancestry (categorical), and income (\$10,000 increments), * p-value <0.05, ** p-value <0.01, *** p-value <0.0001. Note that the model evaluating BMI as a primary risk factor did not also adjust for BMI as a covariate. | | |

Combining the most common risk factors into a single logistic regression model (obesity, type 2 diabetes, fatty liver disease, and high blood pressure), the most significant risk factor for hospitalization remained obesity (defined as BMI > 30), which accounted for a doubling in the risk of hospitalization (aOR = 2.07, 95% CI = 1.66 - 2.58) after adjusting for age, sex, ancestry, education, and household income and the other conditions (Table 3). In this model, African Americans were 83% more likely to be hospitalized for COVID-19 (aOR = 1.83, 95% CI 1.33 - 2.52). Socio-economic status was inversely associated with hospitalization risk, with a 4% decrease in hospitalization per $10,000 increase in median income in the zip code of residence. High school or less education conferred a 38% increased risk in hospitalization (aOR = 1.39, 95% CI 1.10 - 1.74).

**Table 3:** Adjusted odds ratios and 95% confidence intervals from a multivariate logistic regression model estimating the relationship between socio-demographic and preexisting health conditions on COVID-19 hospitalization among all test positive cases.

| Model variable | Adjusted OR | 95% confidence interval |
| --- | --- | --- |
| Sex (female) | 0.79 | 0.68 - 0.93** |
| Age (10-yr increase) | 1.39 | 1.31 - 1.46*** |
| **Socio-economic status** |  |  |
| HH income zip code (\$10k increase) | 0.96 | 0.94 - 0.99** |
| High school or less education | 1.38 | 1.10 - 1.74** |
| **Body Mass Index** |  |  |
| Underweight BMI (19.9 or less vs. normal) | 1.24 | 0.76 - 2.03 |
| Overweight BMI (24.9-29.9 vs. normal) | 1.27 | 1.01 - 1.60* |
| Obese BMI (30+ vs. normal) | 2.07 | 1.66 - 2.58*** |
| **Race/ ethnicity** |  |  |
| Latino vs. European | 1.24 | 1.01 - 1.52* |
| Other non-European vs. European | 1.38 | 1.00 - 1.89* |
| African American or Black vs. European | 1.83 | 1.33 - 2.52*** |
| **Pre-existing conditions** |  |  |
| High blood pressure | 1.29 | 1.08 - 1.54** |
| Type 2 diabetes | 1.48 | 1.15 - 1.91** |
| Fatty liver disease | 1.61 | 1.24 - 2.10*** |
| All variables shown were in the model predicting hospitalization out of all those with a COVID-19 positive test. * p-value <0.05, ** p-value <0.01, *** p-value <0.001 |  |  |

### Geographic distribution of cases

The prevalence of infection, estimated as the number of reported test positives in a state relative to the number of study participants in the state, varied across geographic regions. The highest proportions of positive COVID-19 tests were reported in New York (4.4%) and New Jersey (3.3%), and the lowest proportions were reported in Maine (0.4%) and West Virginia (0.4%). As the majority of case data was collected between late April and early June, hotspots that developed earlier in the pandemic are better represented compared to those that arose later in the course of the pandemic (Figure 3a; Supplementary Figure 1). Nonetheless, the self-reported prevalence of positive COVID-19 tests at the US state level was reasonably well correlated with the number of positive tests reported per capita^16^ as of July (Figure 3b; Pearson r = 0.85). However, the prevalence of self-reported COVID-19 test positive status was higher in the 23andMe database than per-capita estimates, likely reflecting differences in the composition of the 23andMe database and the general population, and potential selection bias arising from individuals with a positive test potentially being more likely to choose to participate in the study.

**Figure 3:**
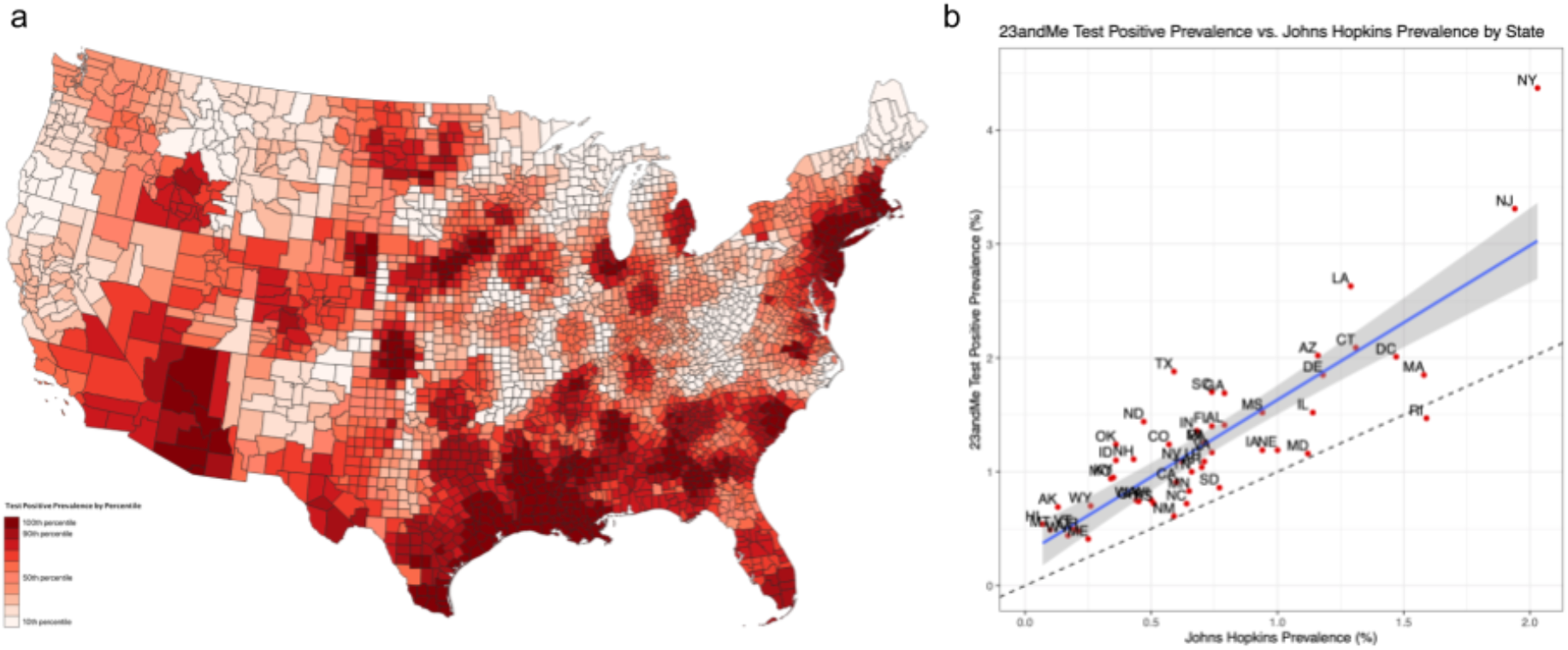
a) Proportion of test positive COVID-19 study respondents in the continental United States combining all reported cases from February - July 2020. The map has been smoothed using a weighted local sum for each county combining data from 15 neighboring counties and weighting by sample size. b) Scatter plot of state-level COVID-19 test positive prevalence as assessed in the 23andMe database compared to that obtained from national statistics^16^.

### GWAS

Across the 5 phenotypes, the trans-ethnic meta-analysis identified 11 genome-wide significant associations from 7 distinct regions of the genome (Figure 4; Supplementary Figures 2 - 4; Supplementary Table 5).

**ABO:** In our phenotype contrasting COVID-19 test positive individuals to test negative individuals, we identified an association at chr9q34.2, with index SNP rs9411378 (p-value = 5.3e-20, C allele OR = 0.857; Figure 5). This index SNP is in LD with a functional variant in the ABO gene, specifically rs8176719 (r^2^ = 0.57 in the European population), which is a well-known single-nucleotide deletion that usually confers a type O blood group when present in the homozygous form. While multiple rare variants elsewhere within the *ABO* gene can contribute to blood group determination, individuals heterozygous for the deletion are most likely to have blood groups A or B, whereas individuals without any copies of the deletion are most likely to have blood groups A, B, or AB.

**Figure 4:**
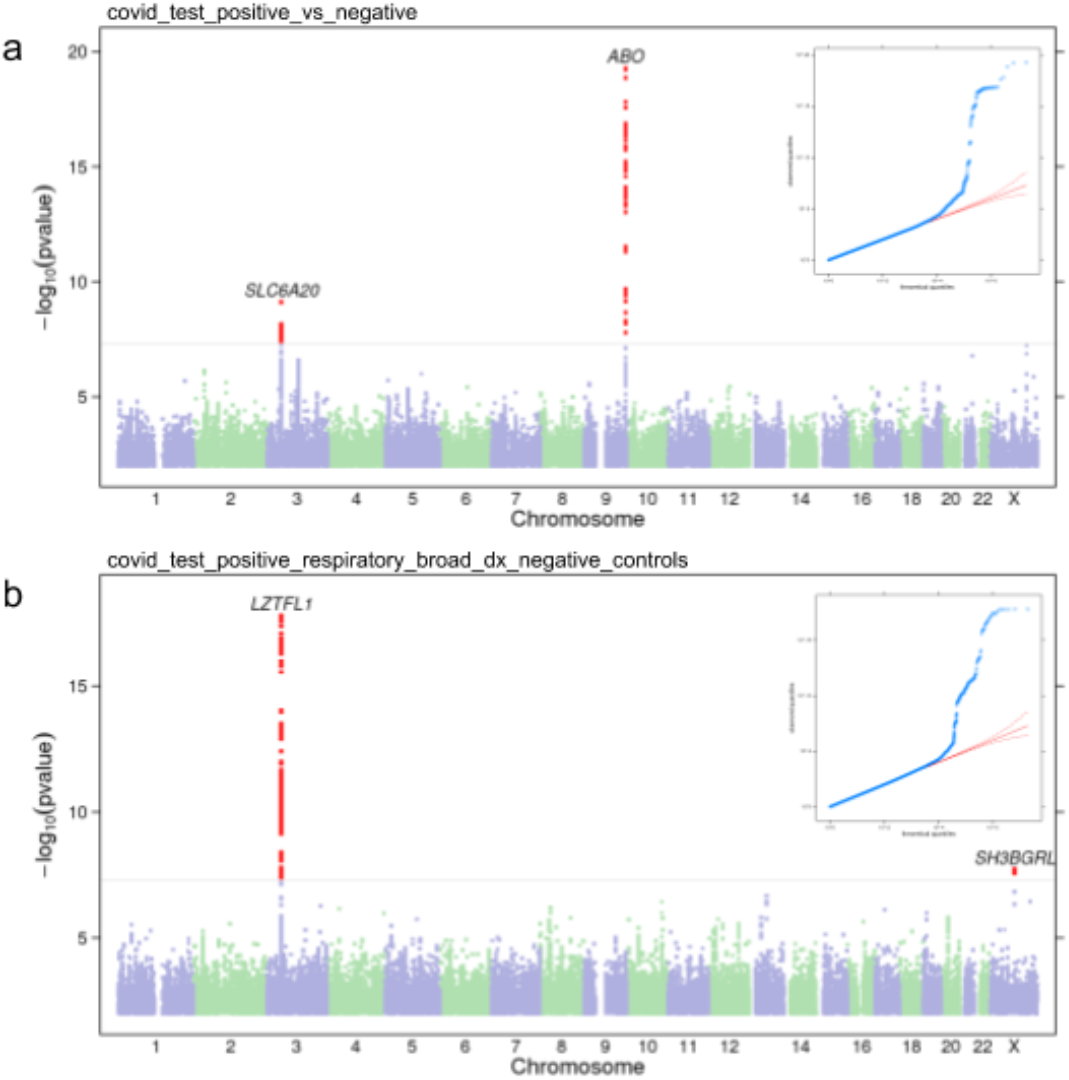
Manhattan and QQ plots for ‘COVID-19 test +ve vs COVID-19 test −ve’ a) and ‘COVID-19 severe respiratory symptoms’ b) from trans-ethnic meta-analysis. The nearest gene to the index SNP is indicated above each association peak.

**Figure 5:**
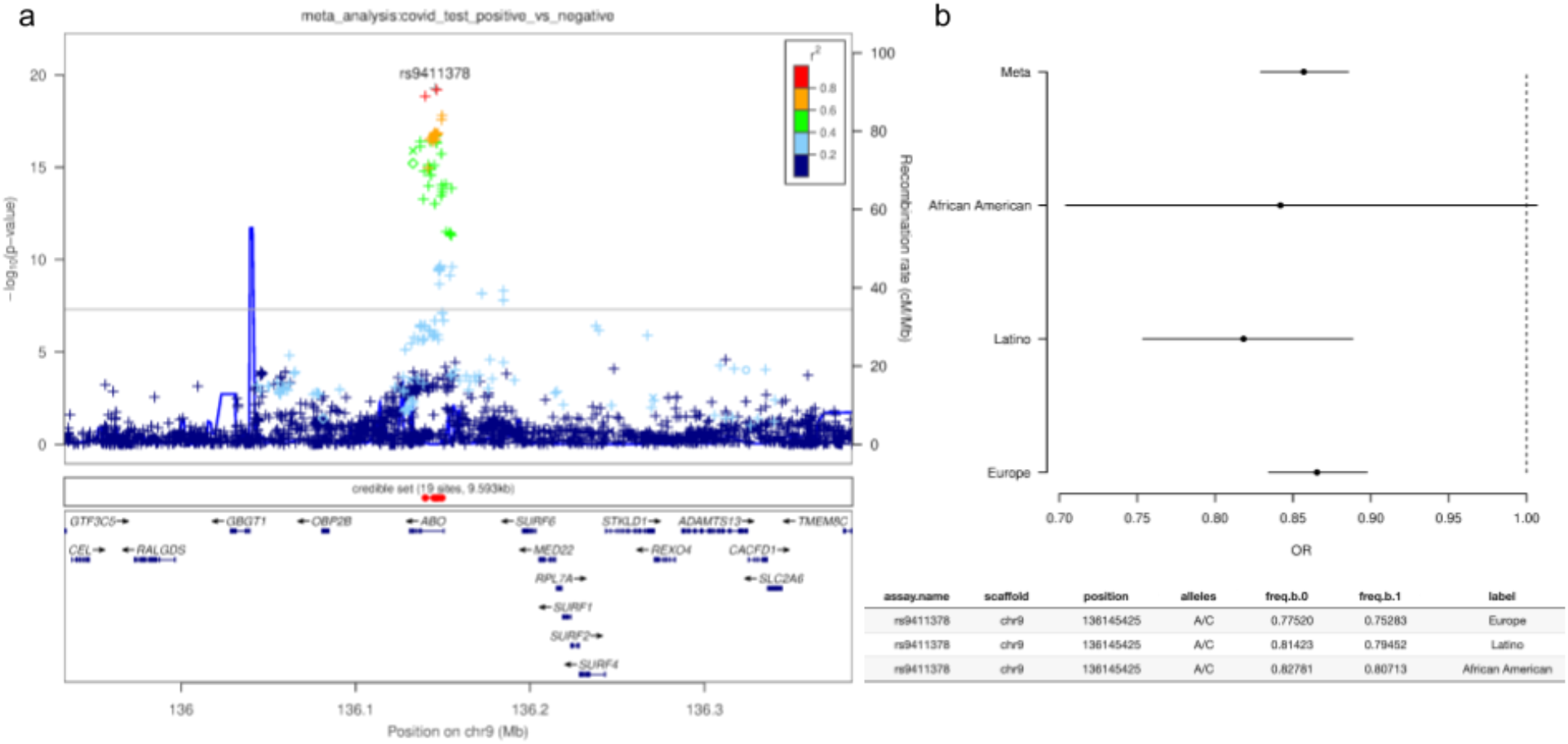
a) Regional plot around the *ABO* locus for phenotype ‘COVID-19 test positive vs COVID-19 test negative’ from trans-ethnic meta-analysis. Imputed variants are indicated with ‘+’ symbols or ‘x’ symbols for coding variants. Where imputed variants weren’t available, genotyped variants are indicated by ‘o’ symbols or diamond symbols for coding variants. b) Odds ratios and 95% confidence intervals for each population.

To further understand the relationship between COVID-19 test positive status and ABO blood group, we used genetically determined blood group assignments (see Methods) and estimated the contribution to risk by comparing each blood group against each of the others. For all COVID-19 phenotypes, we found that the O blood group was protective when compared to other blood groups, whereas blood groups A, B, and AB did not differ from each other (Figure 6; Supplementary Figure 5, Supplementary Table 6). Direction of effect was broadly consistent across populations (Supplementary Figure 6). We see a similar size of effect in both the diagnosis and severity phenotypes, albeit with the severity phenotypes not achieving statistical significance, possibly due to smaller sample size. We also note that the association with COVID-19 diagnosis is in contrast to the ABO results obtained when considering individuals that reported influenza symptoms in the years before the COVID19 pandemic, where blood group O appears to be a risk factor (see Discussion).

There have been preliminary reports suggesting that the rhesus factor (Rh) can also contribute to differences in susceptibility and severity^17^. We do not detect a genetic association at the RHD locus, which suggests that Rh is not a major risk factor by itself independent of ABO blood group. In order to further investigate, we also compared positive and negative forms of each ABO blood group, and detected no significant difference in any comparison (Supplementary Table 7).

**Figure 6:**
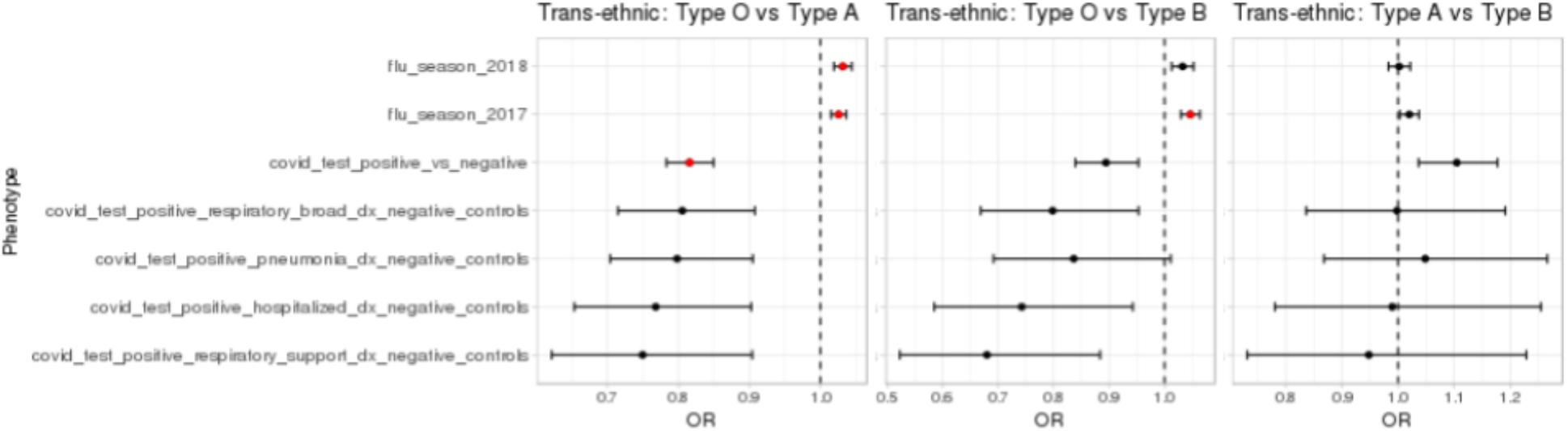
Comparison of blood groups across phenotypes. Statistically significant associations (p-value < 0.001) are highlighted in red. Blood group AB did not show differentiation from groups A or B (Supplementary Figure 5).

**chr3p21.31:** We identified an association at chr3p21.31, which was shared across all phenotypes (Figure 7a; Supplementary Figure 7). The association appeared strongest in our phenotypes related to respiratory symptoms, with the lowest p-value observed in the severe respiratory symptoms phenotype (index SNP rs13078854, alleles A/G, p-value = 1.6e-18), and with a relatively large estimated effect size (G allele OR = 0.592, 95% CI 0.527 - 0.665). The majority of support for this association comes from the European population (Figure 7b), likely reflecting the larger sample size for this cohort. However, the risk allele is also more common in the European population, with the rs13078854 A allele having frequency 7.8%, 5.8%, and 2.7% in the European, Latino, and African American populations, respectively.

The credible set for this locus overlaps the *LZTFL1* gene, although none of the variants in the credible set alter the resulting protein. The locus also contains other nearby genes that could plausibly be driving the association, including *SLC6A20, CCR9, FYCO1, CXCR6*, and *XCR1*. In our data, the index SNP is in high LD (r^2^ > = 0.85) with eQTLs for *SLC6A20*, as detected in GTEx within breast epithelium and esophagus muscularis mucosa^17^, suggesting increased expression of *SLC6A20* correlates with increased risk of severe outcomes.

**Figure 7:**
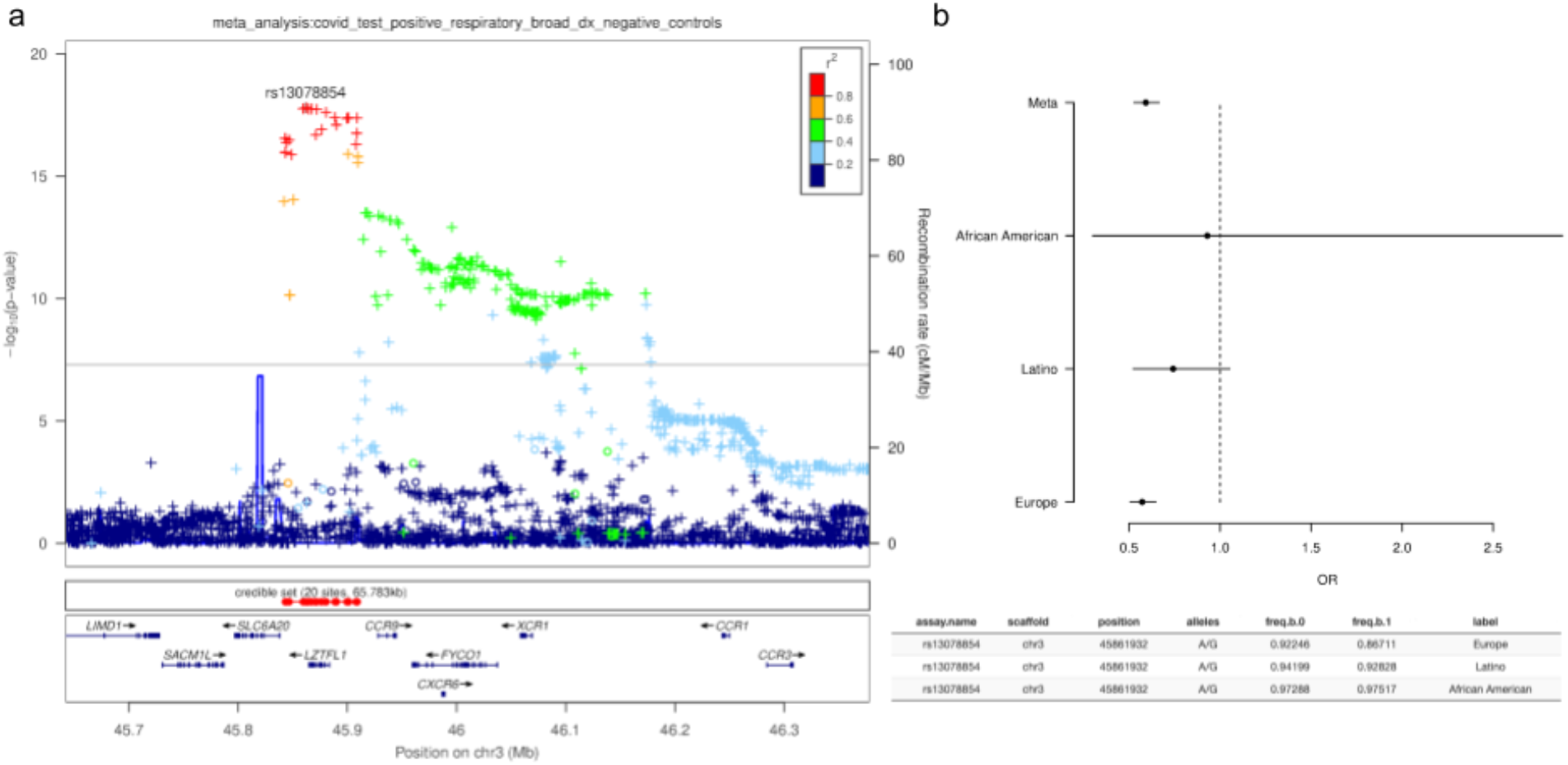
a) Regional plot around the chr3 locus for the COVID-19 severe respiratory symptoms phenotype. Imputed variants are indicated with ‘+’ symbols or ‘x’ symbols for coding variants. Where imputed variants weren’t available, genotyped variants are indicated by ‘o’ symbols or diamond symbols for coding variants. b) Odds ratio estimate and 95% confidence interval for each population.

Given the reported differences in outcome severity between males and females, we tested for a difference in effect at the chr3p21.31 locus. Testing rs13078854 separately in males and females for the severe respiratory symptoms phenotype gave an OR = 0.49 (95% CI: 0.41 - 0.59) in males and OR = 0.69 (95% CI: 0.58 - 0.82) in females, with the difference being moderately significant (p-value = 0.003; z-test). On the basis of the association between ABO and COVID-19, we further hypothesized that the chr3p21.31 locus may show a difference in effect size depending on ABO blood type. Conditioning on blood type O, we estimated the OR of rs13078854 to be 0.63 (95% CI: 0.52 - 0.77), whereas conditioning on any other blood type gave an OR of 0.57 (95% CI: 0.49 - 0.66). We therefore conclude that ABO blood type does not modulate the effect at the chr3p21.31 locus (p-value = 0.80; z-test).

#### Other associations

In addition to the two main associations, we observed 5 weaker associations which, while achieving genome-wide significance, typically only include a small number of low-frequency variants within the association peak, and may represent false-positive associations (Supplementary Table 5).

## Discussion

The COVID-19 pandemic represents a unique emergency in recent human history, and has dramatically accelerated the pace of scientific investigation into the effects of the virus on human health. In this paper, we have utilized a direct-to-consumer research platform to collect data regarding experiences of COVID-19 at large scale and over a compressed timeline.

Both Latino and African American groups reported a higher rate of COVID-19 infection and, as reported elsewhere^2,18,19^, our data show an elevated risk of hospitalization in these populations. For Latino respondents, the higher rate of hospitalization was broadly consistent with the higher rate of infection. However, for African American respondents, the risk of hospitalization was disproportionately high, and remained so after adjusting for socio-demographic characteristics, age, sex, obesity, type 2 diabetes, hypertension, and fatty liver disease.

Our data also strengthens the evidence for a role for ABO in COVID-19 host genetics. ABO blood group has been reported as a risk factor for both COVID-19 susceptibility^20^ and severity^15^, and is notable given the reported links between COVID-19 and blood clotting complications^21,22^. Our data supports a role in susceptibility to infection, suggesting that blood group O is protective in contrast to non-O blood groups. Whereas previous reports suggested protection was limited to the rhesus positive group^23^, our data does not support that conclusion.

The mechanism by which ABO is associated with COVID-19 is unclear, but ABO blood groups can play a direct role in pathogen infection by serving as receptors and/or coreceptors^24^. SARSCoV-2 is an enveloped virus that carries ABO antigens on the viral spike (S) glycoprotein and host envelope glycolipids. Recent work has shown the SARS-CoV-2 S protein interacts with multiple host C-type lectin receptors in a glycosylation-dependent manner^25,26^, similar to previous work on the SARS-CoV virus from the earlier SARS outbreak^27,28^. Differential glycosylation of the spike protein or the envelope glycolipids from expression of different ABO glycosyltransferases may then impact the binding and internalization of SARS-CoV-2 viral particles. Others have speculated that the lower susceptibility of blood group O could be linked to anti-A blood antibodies inhibiting the adhesion of coronavirus to ACE2-expressing cells, thereby providing protection^29^.

The ABO locus is also highly pleiotropic^30^ and exhibits complex population structure^31^. Interestingly, while old literature regarding the association between ABO and influenza is inconsistent^32^, our own data suggests that blood type O is actually a risk factor for seasonal flu. This is notable because COVID-19 testing in the United States was largely restricted to individuals with flu-like symptoms at the time we were collecting the majority of our data. As such, it is possible that the population of individuals receiving COVID-19 tests was enriched for influenza cases, and the apparent protective nature of ABO for COVID-19 could arise from a subtle form of collider bias. However, our data also shows that blood type O is protective in the severe-outcome population, which wouldn’t be expected under the collider bias hypothesis.

Likewise, our data strengthens the evidence of association at the chr3p21.31 gene cluster, first identified by Ellinghaus *et al*.^15^. The locus contains multiple genes (*SLC6A20, LZFTL1, CCR9, CXCR6, XCR1, FYCO1*) that could be functionally implicated in COVID-19 pathology. In particular, *SLC6A20* can be linked to the association via eQTLs in relevant tissue and has been noted^15^ as potentially forming a complex with angiotensin converting enzyme 2 (ACE2), the cell surface receptor for SARS-CoV-2 viral entry^33,34^. It is possible that increased *SLC6A20* expression leads to increased ACE2 protein levels and greater viral uptake. *LZFTL1* has been implicated in ciliogenesis and intracellular trafficking of ciliary proteins, which may impact airway epithelial cell function. As noted elsewhere^15^, CXCR6 promotes NKT cell and tissue resident memory CD8+ T cells residence in the lung^35^ and plays a role in the trafficking of T lymphocytes to the bronchial epithelia during respiratory infection and inflammatory lung disease. CCR9 predominantly regulates T cell homing to the gut, which may indirectly impact the response in the lung, however it has also been shown to regulate eosinophil recruitment to the lung^36^. Recent studies have identified elevated chemokines and eosinophilia as a hallmark of severe disease^37–39^, but additional work will be required to define any functional contribution of these genes to the genetic association with COVID-19.

In phenotypes contrasting individuals with severe COVID-19 symptoms to controls without a COVID-19 diagnosis, the risk variants at the chr3p21.31 locus achieve odds ratios of approximately 2.0 in our data, which is relatively large in the context of GWAS studies. Given the risk alleles are also relatively common (∼3 - 8% frequency, depending on population), it is likely that this locus makes a meaningful contribution to determining why some individuals experience severe COVID-19 outcomes. However, while the population sample sizes in our study differ considerably, we found little evidence to suggest that allele frequency differences at this locus could account for the higher rate of severe outcomes from COVID-19 for non-European ancestry groups. In fact, the primary risk allele at the chr3p21.31 locus is most common in European populations, and less common in Latino, and African American populations.

This study is a testament to the power of the 23andMe research platform, which in less than four months enabled over one million research participants to contribute to the study of a novel disease. However, there are notable caveats to relying on self-reported data for a disease with lethal outcomes. Namely, the cases identified in this study were healthy enough to respond to the survey therefore are likely biased towards a healthier case population than otherwise exists. In addition, 23andMe research participants are a self-selected group and may not reflect the general population. Furthermore, the scarcity of testing has likely further obscured the true picture of SARS-CoV-2 infections in the United States, leading to misclassification of true cases as controls in this study if they did not receive a positive test result. The effect of these types of error would bias the reported effect estimates towards the null, meaning that the true impact of risk factors reported here may be expected to be larger if the sample were randomly drawn from the broader population and had perfect case and control classification.

## Data availability

The full set of de-identified summary statistics can be made available to qualified investigators who enter into an agreement with 23andMe that protects participant confidentiality. Interested investigators should visit the following: https://research.23andme.com/covid19-dataset-access/.

## Data Availability

https://research.23andme.com/covid19-dataset-access

## Acknowledgements

We thank the 23andMe research participants who made this study possible. We would also like to thank Altovise Ewing, Aaron Petrakovitz, Anne Park, Anne Silk, Aushawna Collins, Becky Macintosh, Carolyn Kao, Courtney Ball, Christine Pai, David Hinds, Devyn Parry, Elo Ratcliff, Emily Bullis, Eric Hall, Farwa Alam, Jacquie Haggarty, Jess Christenson, Jim Lawrence, Jimmy Chau, Josie Shaw, Joe Cackler, Karl Heilbron, Katelyn Kukar, Katie Watson, Marianna Frendo, Olivia Valenti, Ryan Workman, Rachel Lopatin, Robert Bell, Rose Eckert, Sam Rodgers, Sarah Rys, Shawna Averbeck, Shirin Fuller, Vanessa Lane, and Yunxuan Jiang for contributions and insights.

We also thank the 23andMe Research Team: Barry Hicks, Chao Tian, Devika Dhamija, Elizabeth Babalola, Elizabeth S. Noblin, Ethan M. Jewett, G. David Poznik, Gabriel Cuellar Partida, Jared O’Connell, Jingchunzi Shi, Joanna L. Mountain, Joyce Y. Tung, Katarzyna Bryc, Karen E. Huber, Keng-Han Lin, Kimberly F. McManus, Kipper Fletez-Brant, Marie K. Luff, Matthew H. McIntyre, Maya Lowe, Meghan E. Moreno, Peter Wilton, Pierre Fontanillas, Priyanka Nandakumar, Sahar V. Mozaffari, Sarah L. Elson, Sayantan Das, Steven J. Micheletti, Suyash Shringarpure, Vinh Tran, Wei Wang, Will Freyman, and Xin Wang.

Members of the 23andMe COVID-19 Team are: Adam Auton, Adrian Chubb, Alison Fitch, Alison Kung, Amanda Altman, Andy Kill, Anjali Shastri, Catherine Weldon, Chelsea Ye, Daniella Coker, Janie Shelton, Jason Tan, Jeff Pollard, Jennifer McCreight, Jess Bielenberg, John Matthews, Johnny Lee, Lindsey Tran, Michelle Agee, Monica Royce, Nate Tang, Pooja Gandhi, Raffaello d’Amore, Ruth Tennen, Scott Dvorak, Scott Hadly, Stella Aslibekyan, Sungmin Park, Taylor Morrow, Teresa Filshtein Sonmez, Trung Le, and Yiwen Zheng.

## Supplementary Material

Supplementary Figure 1: Estimated prevalence of COVID-19 test positive individuals on a per-county basis within the continental United States.

Supplementary Figure 2: Manhattan and QQ plots for ‘COVID-19 test positive with hospitalization’ phenotype from trans-ethnic meta-analysis. Cases reported a positive COVID-19 test and were hospitalized with their symptoms. Controls did not report a COVID-19 diagnosis (were neither diagnosed with nor tested positive for COVID-19).

Supplementary Figure 3: Manhattan and QQ plots for ‘COVID-19 test positive with respiratory support’ from trans-ethnic meta-analysis. Cases reported a positive COVID-19 test and received respiratory support in the form of supplementary oxygen or ventilation. Controls did not report a COVID-19 diagnosis (were neither diagnosed with nor tested positive for COVID-19).

Supplementary Figure 4: Manhattan and QQ plots for ‘COVID-19 test positive with pneumonia’ from trans-ethnic meta-analysis. Cases reported a positive COVID-19 test and experienced pneumonia. Controls did not report a COVID-19 diagnosis (were neither diagnosed with nor tested positive for COVID-19).

Supplementary Figure 5: Comparison of AB blood group to other ABO blood groups in the trans-ethnic meta-analysis. Statistically significant associations (p-value < 0.001) are highlighted in red.

Supplementary Figure 6: Comparison of blood groups across phenotypes and across populations: European, African American, and Latino. Statistically significant associations (pvalue < 0.001) are highlighted in red.

Supplementary Figure 7: Forest plot of chr3p21.31 index SNP (rs13078854) in the trans-ethnic analysis and for each population. Genome-wide significant associations are highlighted in red.

Supplementary Table 1: Geo-targeted send of COVID-19 study emails to 23andMe customers between April and June 2020.

Supplementary Table 2: GWAS phenotype definitions.

Supplementary Table 3: ABO assignments derived from haplotypes defined by 3 SNPs.

Supplementary Table 4: Precision and Recall of ABO haplotype model, compared to self-reported ABO blood group.

Supplementary Table 5: Genome-wide significant association index SNP from the trans-ethnic meta analysis.

Supplementary Table 6: Pairwise comparisons of ABO blood groups.

Supplementary Table 7: Comparison of rhesus factors within ABO blood groups.

